# Increasing prevalence of HIV-1 Transmitted Drug Resistance in Portugal: implications for first line treatment recommendations

**DOI:** 10.1101/2020.03.17.20033092

**Authors:** Marta Pingarilho, Victor Pimentel, Isabel Diogo, Sandra Fernandes, Mafalda Miranda, Andrea Pineda-Pena, Pieter Libin, Kristof Theys, M. Rosário O. Martins, Anne-Mieke Vandamme, Ricardo Camacho, Perpétua Gomes, Ana Abecasis, on behalf of the Portuguese HIV-1 Resistance Study Group

## Abstract

**Background:** Treatment for all recommendations has allowed access to antiretroviral (ARV) treatment to an increasing number of patients. This minimizes transmission of infection but can potentiate the risk for development of transmitted drug resistance (TDR) and acquired drug resistance (ADR).

**Objective:** To study the trends of TDR and ADR in patients followed in Portuguese hospitals between 2001 and 2017.

**Method:** 11911 patients of the Portuguese REGA database were included. TDR was defined as the presence of one or more surveillance drug resistance mutations according to the WHO surveillance list. Phenotypic resistance to ARV was evaluated with Standford HIVdb v7.0. Patterns of TDR, ADR and prevalence of mutations over time were analysed with logistic regression.

**Results:** The prevalence of TDR increased from 7.9% in 2003 to 13.1% in 2017 (p_for-trend_<0.001). This was due to a significant increase of both resistance mutations to nucleotide reverse transcriptase inhibitors (NRTIs) and non-nucleotide reverse transcriptase inhibitors (NNRTIs) from 5.6% to 6.7% (p_for-trend_=0.002) and 2.9% to 8.9% (p_for-trend_<0.001), respectively. TDR to Protease Inhibitors decreased from 4.0% in 2003 to 2.2 in 2017 (p_for-trend_ =0.985). Paradoxically, the prevalence of ADR declined from 86.6% in 2001 to 51.0% in 2017 (p_for-trend_<0.001) caused by a declining drug resistance to all ARV classes (p_for-trend_<0.001).

**Conclusions:** While ADR is declining since 2001, TDR has been increasing, reaching a value of 13.1% by the end of 2017. It is urgent to develop public health programs to monitor levels and patterns of TDR in newly diagnosed patients.

## Introduction

In 2014, the WHO proposed the 90-90-90, an ambitious treatment target to help end the AIDS pandemic. In Portugal, a national report estimated that in 2017, 38.959 people were living with HIV, with an incidence of 8,6 cases per 100.000 habitants in 2017, one of the highest in Europe. Of these, 91.7% of patients were diagnosed, 86.8% of the diagnosed were receiving antiretroviral treatment (ART) and 90.3% of the treated patients (TP) were virally suppressed [1]. In summary, Portugal is very close to achieving the 90-90-90 objectives, lacking only a small proportion of patients under treatment.

A major threat to achieve the 90-90-90 target is ARV drug resistance. While ART largely decreases HIV-related morbidity and mortality, improves quality of life of HIV-1 infected patients and reduces the risk of onward transmission [2–4], its scale up can potentiate the risk for development of ARV drug resistance.

International guidelines consistently recommend that newly diagnosed individuals should be tested for ARV drug resistance to detect potential transmitted drug resistance (TDR) and guide the selection of ART regimens [5,6]. This procedure minimizes the risk of experiencing virologic failure after starting ART due to the selection of resistant HIV isolates, resulting in acquired drug resistance (ADR).

Active surveillance of TDR and ADR is crucial to understand factors involved in the transmission of HIV-1 drug resistance and, also, to help to design effective ART treatment guidelines in different epidemic settings. Drug resistance evolves dynamically and therefore it is extremely important to monitor temporal trends of TDR and ADR.

In this study, we aim to describe the temporal trends of TDR and ADR between 2001 and 2017, as well as the single mutations involved in that resistance and to identify predictors of TDR among HIV-1 infected patients treated in Portuguese hospitals.

## Methods

### Study population

The protocol was in accordance with the declaration of Helsinki and approved by the Ethical Committee of Centro Hospitalar de Lisboa Ocidental (108/CES-2014). The Portuguese HIV-1 resistance database contains anonymized patients’ information, including demographic, clinical and genotype resistance data from patients followed in 22 hospitals located around the country and collected between May 2001 and December 2017. All patient data collected in RegaDB [7] were generated during routine clinical care. Individuals aged 18 years or over and which had a HIV-1 drug resistance test performed were included. Patients’ viral sequences were categorized into drug naïve (DN) and treated patients (TP). The CD4 cell count and HIV-1 viral load measured closest to the date of the resistance test were included in the analysis.

### Drug resistance analyses and subtyping

The genomic data included protease and reverse transcriptase sequences obtained through population sequencing completed in Laboratório de Biologia Molecular do Hospital de Egas Moniz, CHLO. TDR was defined as the presence of one or more surveillance drug resistance mutations (SDRM) according to the WHO 2009 surveillance list [8]. Nucleotide sequences were submitted to the Calibrated Population Resistance tool - HIV drug resistance database version 6.0. Phenotypic resistance to ARV drugs was evaluated using the Standford HIVdb v8.4. Using this algorithm, ADR mutations were also identified. HIV-1 subtypes and circulating recombinant forms (CRF) were determined as previously described [9,10].

### Statistical analysis

Proportions and confidence intervals for proportions were calculated using 95% Wilson confidence interval for binomially distributed data. The differences between the prevalence of resistance in naïve and treated patients were analyzed using the Mann-Whitney-U-test and the X^2^ tests. Logistic regression was used to examine the association between demographic and clinical factors and the prevalence and the impact of SDRMs and to analyse trends over time. Results were considered statistically significant when p-values were below 0.05. All analyses were conducted in SPSS Statistic version 25 software and R3.5.1. More information in Supplementary File 1.

## Results

### Population

Of 12792 patients, 11912 (93,1%) patients presented viral sequences. More than a half (7310, 61.4%) were from drug naïve and 3848 (32.3%) from treated patients. ART status was not available for 754 (6.3%) patients. Population characteristics are described in text and in Table A in Supplementary File 1.

### Transmitted HIV drug resistance (TDR)

Overall, the prevalence of TDR between 2001 and 2017 was 9.4%. Non-nucleoside reverse transcriptase inhibitor (NNRTI) mutations were detected in 5.0% of drug-naive patients, 4.0% for nucleoside reverse transcriptase inhibitor (NRTI) and 2.8% for protease inhibitors (PI). 8.5% presented single class resistance, 0.7% dual class resistance and 0.2% triple class resistance (Table 1).

**Table 1.**
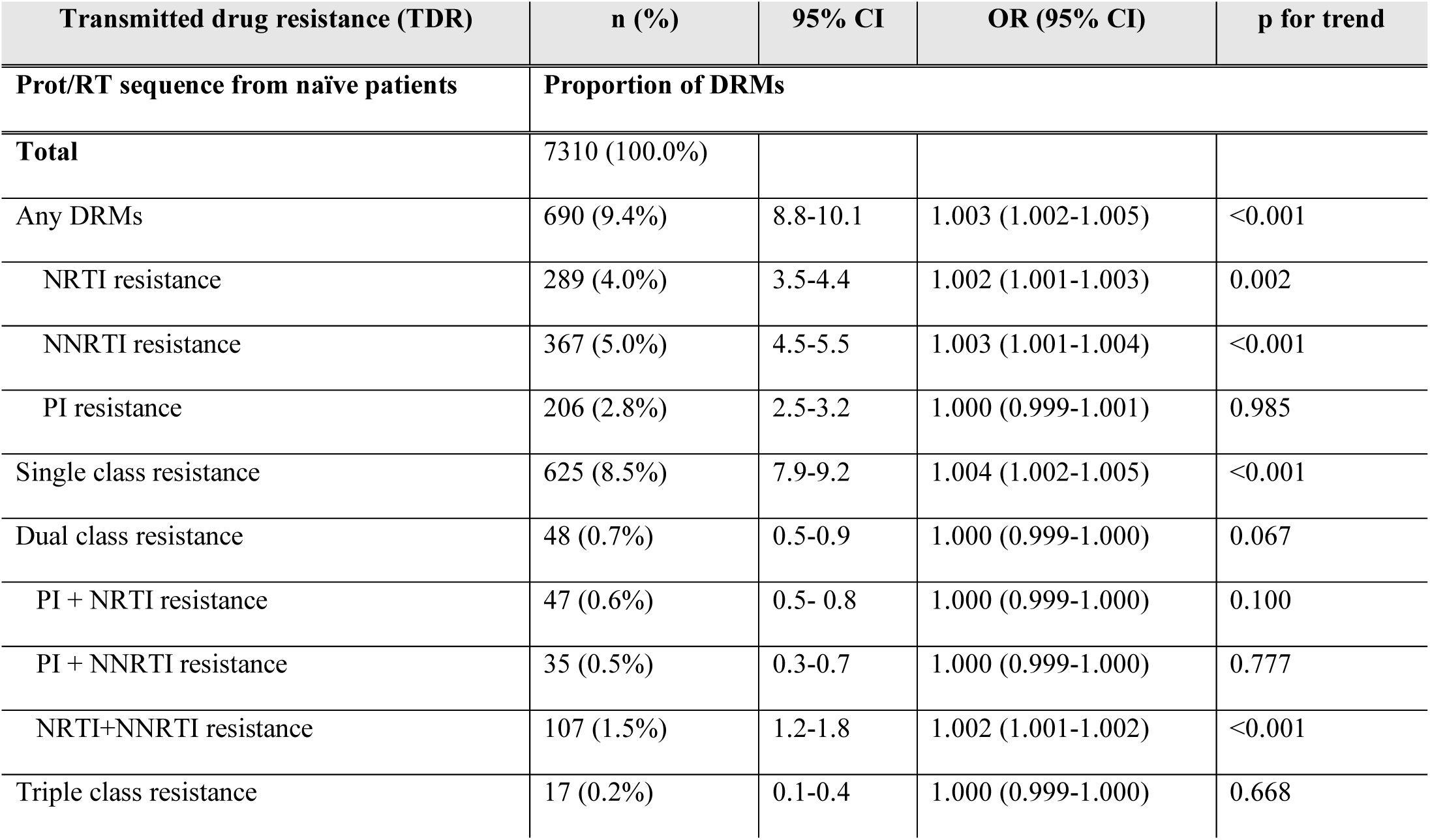
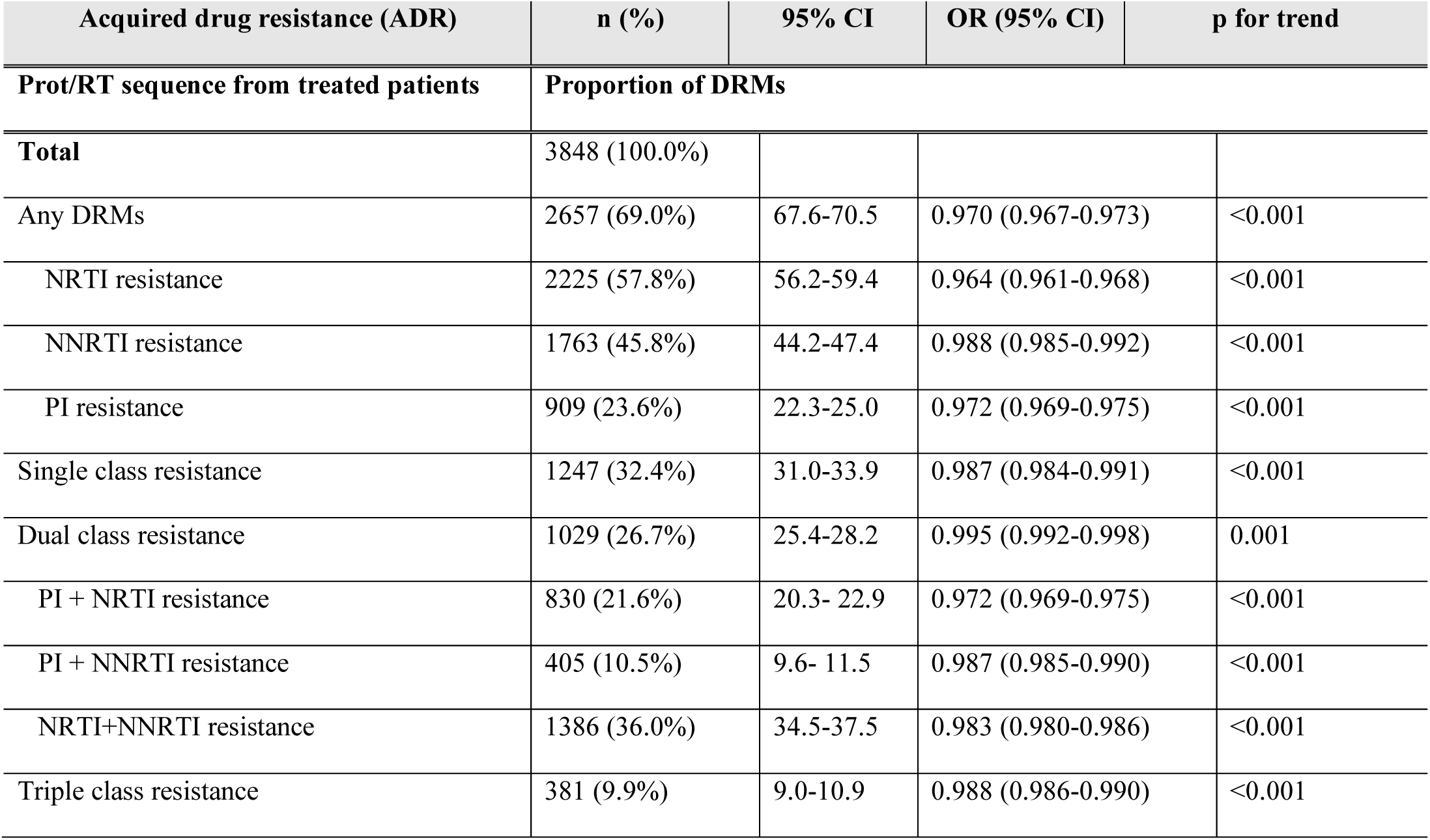
Proportion of transmitted drug resistance between 2003 and 2017 and of acquired drug resistance between 2001 and 2017

Trends for TDR were determined only between the years 2003 and 2017, since between 2001 and 2002 the number of patients tested for ARV drug resistance was not large enough to allow for robustness in statistical analyses.

TDR presented a significantly increasing trend from 7.9% in 2003 to 13.1% in 2017 (p_for- trend_<0.001). This trend is steeper in the last four years analysed (2014 to 2017) (p_for-trend_=0.008). Since treatment for all recommendations were implemented in Portugal in September 2015, trends between September 2015 and December 2017 was also analysed, but was not found to be statistically significant (p_for-trend_=0.507) (Figure 1(A) and Table 1, and Table B in Supplementary File 1).

**Figure 1:**
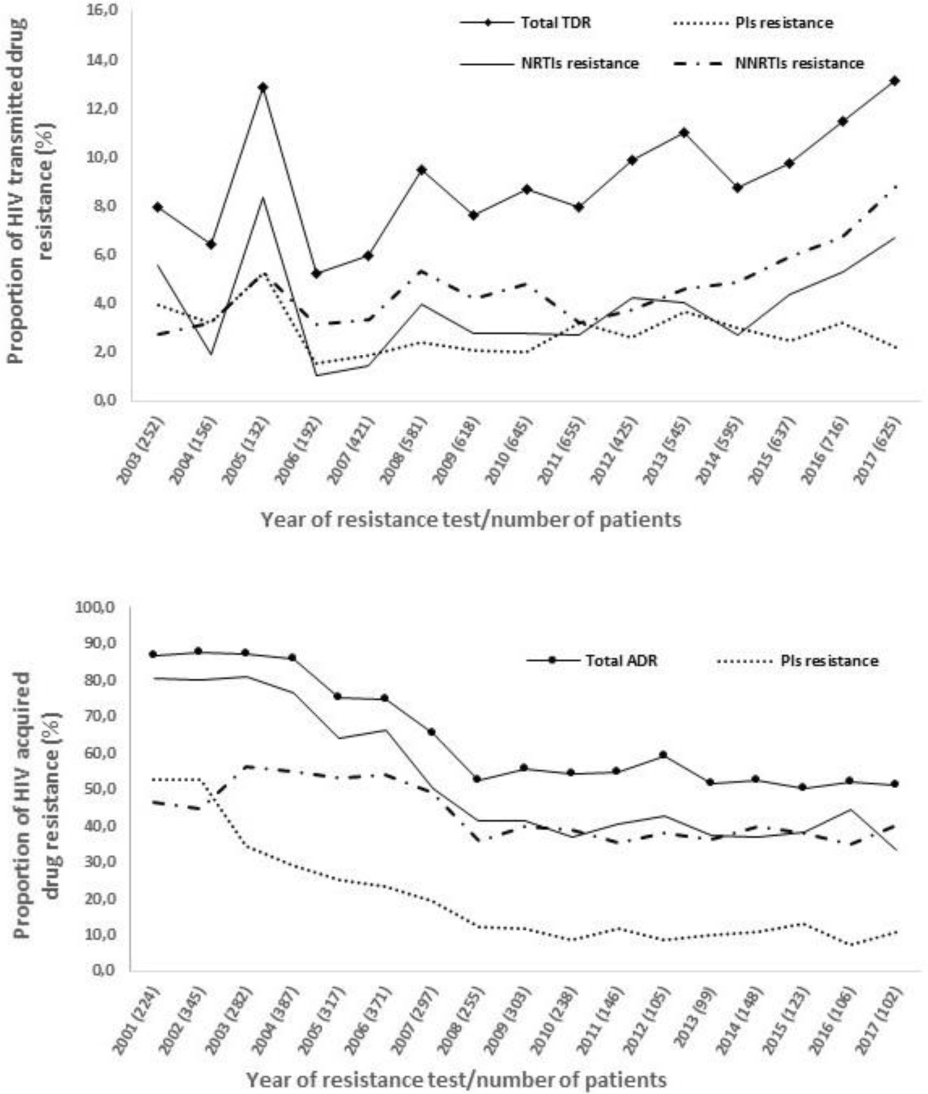
Proportion of (A) transmitted drug resistance (TDR) in sequences from naïve patients between 2003 and 2017 and (B) of acquired drug resistance (ADR) in treated patients between 2001 and 2017.

This increasing trend was significant for two drug classes: NRTIs (5.6% in 2003 to 6.7% in 2017, p_for-trend_= 0.002) and NNRTIs (2.9% in 2003 to 8.9% in 2017, p_for-trend_<0.001). For PIs, on the other hand, there was an opposite trend for lower drug resistance levels in more recent years (4.0% in 2003 to 2.2% in 2017). However, it was not significant (p_for-trend_= 0.985) (Figure 1 (A), Table 1). Single class mutations increased over time from 5.2% (2003) to 12.2% (2017) whereas double class and triple class slightly decreased overtime (p_for-trend_<0.001 for single class; p_for-trend_=0.067 for double class; p_for-trend_=0.668). Double class resistance to NRTIs/NNRTIs combinations presented an increase between 2011 and 2017 (p_for-trend_=0.0604), whereas resistance to combinations of NRTIs/PIs and NNRTIs/PIs just presented a slight increase in the last years (2014-2017) (p_for-trend_=0.272, p_for-trend_=0.072, respectively) (Table 1 and Tables B and C in Supplementary File 1).

The most frequently detected mutations were K103NS (3.2%) conferring resistance to NNRTIs, followed by M41L (1.6%) and M184V/I mutation (1.3%) conferring resistance to NRTIs and L90M (1.2%) conferring resistance to PIs (Supplementary Figure 1(A)).

### Acquired HIV drug resistance (ADR)

ADR prevalence was 69.0%. NRTIs resistance mutations were predominantly identified (57.8%), followed by NNRTIs (45.8%) and PIs (23.6%). 32.4% presented single class resistance, 26.7% dual class resistance and 9.9% triple class resistance (Table 1).

Overall, ADR decreased over time from 86.6 to 50.9 between 2001 and 2017 (p_for-trend_<0.001) and between September 2015 and December 2017 (p_for-trend_=0.836). Resistance to all drug classes presented declining trends over time (2001-2017): resistance to NRTIs dropped from 80.8% to 33.3% (p_for-trend_<0.001); to NNRTIs from 46.4% to 40.2% (p_for-trend_<0.001) and to PIs from 52.7% to 10.8% (p_for trend_<0.001) (Figure 1(B) and Table 1, and Table B in Supplementary File 1).

For single, double and triple class, significant declining trends were also observed: p_for-trend_<0.001 for single class; p_for-trend_=0.001 for double class and; p_for trend_<0.001 for triple class); as well as for combinations of antiretroviral classes. NRTIs/NNRTIs combination decreased from 41.1% to 24.5%, NRTIs/PIs decreased from 51.3% to 8.8% and NNRTIs/PIs decreased from 23.2% to 6.9% between 2001 and 2017 (p_for-trend_<0.001; p_for-trend_<0.001 and p_for-trend_<0.001 for NRTIs/NNRTIs, NRTIs/PIs and NNRTIs/PIs, respectively) (Table 1 and Table C in Supplementary File 1).

The most prevalent mutations conferring resistance to NRTIs in treated patients, were M184IV (45.3%) and TAMs, such as T215YF (17.4%)) and M41L (16.0%). K103NS (26.0%) and L90M (11.3%) were the most prevalent mutations conferring resistance to NNRTIs and to PIs, respectively (Supplementary Figure 1 (B)).

Trends for frequencies of specific drug resistance mutations were also analysed between 2001 and 2017 when mutations had a prevalence greater than 0.5% for drug-naive patients and 5.0% for treated patients. Among drug-naïve patients, although none of the mutations presented a statistically significant trend, we observed an increasing trend for M184V in the last years (Figure 3). For treated patients, on the other hand, PI resistance mutations M46IL, I54VLMTAS, V82ATSF and L90M presented statistically significant declining trends over time (p_for-trend_ <0.001). The same was observed for all mutations analysed for NRTIs regimens (p_for-trend_ <0.001) (Table D in Supplementary File)

**Figure 3:**
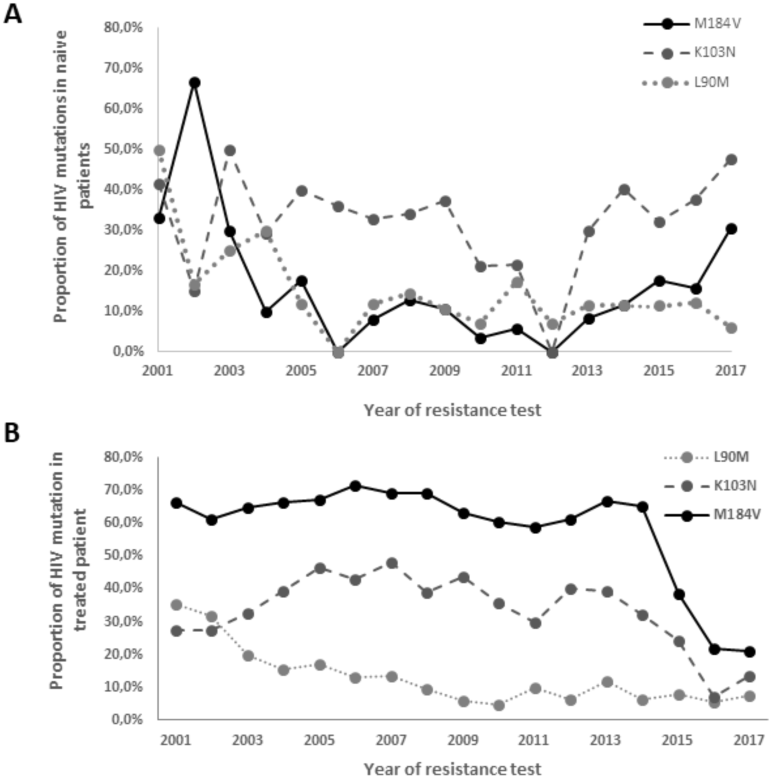
Proportion of M184V, K103N and L90M mutations in (A) drug-naïve patients and (B) treated patients overtime between 2001 and 2017.

### Drug susceptibility

According to the HIVdb Standford database algorithm, 6.6% of DN patients (7310) presented high-level resistance to at least one drug. 5.4% of those have high-level resistance to a drug recommended for first line treatment. NNRTIs presented the highest level of high-level resistance (4.9%), with nevirapine (NVP) having the highest proportion of high-level resistance (4.7%), followed by efavirenz (EFV) with 4.0% of high-level resistance. High-level resistance to NRTIs was 1.7%, with emtricitabine (FTC) and lamivudine (3TC) presenting the highest level (1.3%). High-level resistance to PIs was 1.8%, with atazanavir (ATV) presenting 0.3% of high-level resistance (Figure 2 (A)).

**Figure 2:**
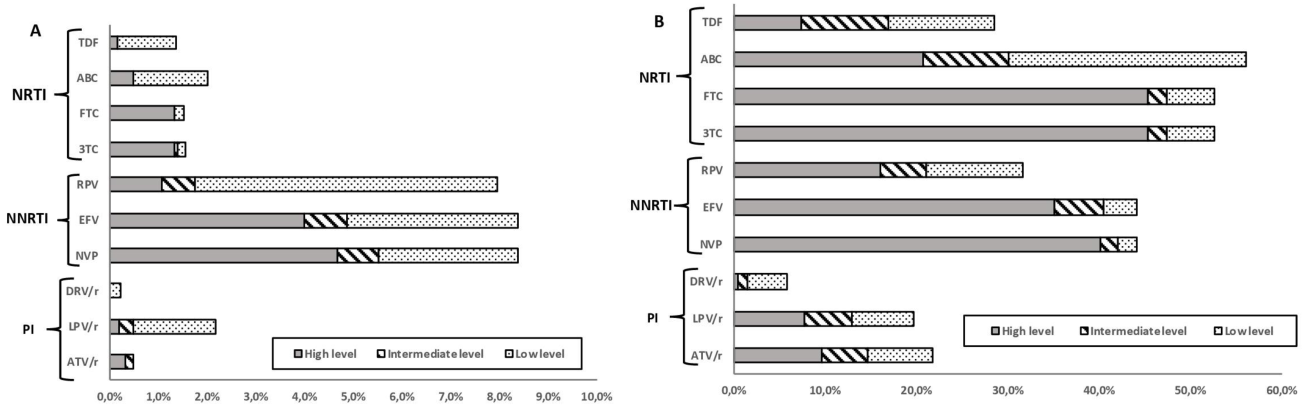
Predicted phenotypic resistance (Standford scores) for antiretroviral drugs currently recommended as first line therapy in Portugal (A) for drug-naives patients (2003-2017) and (B) for treated patients (2003-2017). Scores of low-level (score 2 and 3), intermediate level (score 4) or high-level (score 5) resistance were used to predict phenotypic resistance. Abbreviations: NRTIs, Nucleoside reverse transcriptase inhibitors; NNRTIs, Non-nucleoside reverse transcriptase inhibitors; PIs, Protease inhibitors; FTC, Emtricitabine; TDF, Tenofovir; 3TC, Lamivudine; ABC, Abacavir; EVF, Efavirenz; RPV, Rilpivirine; DRV/r, Darunavir; LPV/r, Lopinavir; ATV/r, Atazanavir

Of 3848 treated patients, (64.0%) presented high-level of resistance to at least one ARV drug class, however considering only the drugs actually used as first line therapy the resistance decreased to 62.1%. For these patients, high-level resistance to NRTIs was the highest (54.4%) with high-level resistance to FTC and 3TC presenting the highest values (45.3%). 40.5% of patients presented high-level resistance to NNRTIs, with 40.1% presenting high-level resistance to NVP and 35% to EFV. High-level resistance to PIs was 19.9%, with ATV presented the highest value (9.6%) (Figure 2 (B)).

### Predictors of TDR

The factors significantly associated with TDR in the univariate model was being infected with subtype B (as compared to non-B subtypes grouped together) and the levels of logVL, while gender presented borderline significance. Although not reaching conventional statistical significance (p<0.05), male gender was associated with higher levels of TDR. It was also observed that being infected with subtype B and logVL above 5.1 presented statistical significance in relation to NRTIs drug resistance; being male, being from sub-Saharan Africa region, being infected with subtype B and logVL above 5.1 presented statistical significance related with PIs drug resistance and in relation to NNRTIs drug resistance logVL above 5.1 was significantly associated (Table E in Supplementary File 1).

Multivariate analysis indicated statistically significant values of any TDR and NRTIs drug resistance for individuals infected with subtype B compared with non-B and presenting logVL higher than 5.1 (Table 2). We further observed that patients with a logVL above 5.1 presented lower levels of K103NS and L90M mutations (p=0.023 and p=0.008, respectively); while patients with a logVL above 4.1 presented lower levels of M184V (p=0.000) (Table 3).

**Table 2.** Multiple regression analysis of factors associated with HIV transmitted drug resistance

| Variable | Any TDR |  | NRTI TDR |  | NNRTI TDR |  | PI TDR |  |
| --- | --- | --- | --- | --- | --- | --- | --- | --- |
|  | OR (95%CI) | p | OR (95%CI) | p | OR (95%CI) | p | OR (95%CI) | p |
| **Gender** |  |  |  |  |  |  |  |  |
| Female * |  |  |  |  |  |  |  |  |
| Male | 1.21 (0.95-1.55) | 0.124 | 1.07 (0.73-1.59) | 0.708 | 1.03 (0.75-1.40) | 0.852 | 1.67 (1.07-2.94) | 0.024 |
| **Age at diagnosis** |  |  |  |  |  |  |  |  |
| 18-25* |  |  |  |  |  |  |  |  |
| 26-40 | 0.97 (0.69-1.36) | 0.860 | 1.33 (0.72-2.44) | 0.361 | 0.75 (0.49-1.15) | 0.186 | 1.78 (0.91-3.50) | 0.093 |
| 41-55 | 0.98 (0.68-1.40) | 0.896 | 1.74 (0.63-3.07) | 0.082 | 0.76 (0.48-1.21) | 0.248 | 1.44 (0.70-2.94) | 0.319 |
| >56 | 1.15 (0.74-1.79) | 0.520 | 1.40 (0.63-3.07) | 0.405 | 1.20 (0.71-2.03) | 0.498 | 2.02 (0.89-4.59) | 0.091 |
| **Subtypes** |  |  |  |  |  |  |  |  |
| B* |  |  |  |  |  |  |  |  |
| Non-B | 0.74 (0.60-0.92) | 0.006 | 0.45 (0.31-0.65) | 0.000 | 1.19 (0.89-1.59) | 0.240 | 0.65 (0.45-0.94) | 0.021 |
| **Viral Load (log<sub>10</sub> copies/ml)** |  |  |  |  |  |  |  |  |
| < 4.0* |  |  |  |  |  |  |  |  |
| 4.1 to 5.0 | 0.73 (0.56-0.96) | 0.024 | 0.42 (0.28-0.63) | 0.000 | 0.70 (0.49-1.00) | 0.054 | 0.76 (0.49-1.19) | 0.234 |
| >5.1 | 0.66 (0.49-0.87) | 0.004 | 0.38 (0.25-0.59) | 0.000 | 0.65 (0.44-0.94) | 0.023 | 0.50 (0.31-0.83) | 0.007 |

**Table 3.** Association for the most prevalent drug-resistance mutation for each major drug class: M184V, L90M, and K103N for NRTI, PI, and NNRTI with viral load

| Variable | M184V |  | K103NS |  | L90M |  |
| --- | --- | --- | --- | --- | --- | --- |
|  | OR (95%CI) | p | OR (95%CI) | p | OR (95%CI) | p |
| **Viral Load (log<sub>10</sub> copies/ml)** |  |  |  |  |  |  |
| < 4.0* |  |  |  |  |  |  |
| 4.1 to 5.0 | 0.30 (0.17-0.54) | 0.000 | 0.76 (0.51-1.12) | 0.164 | 0.64 (0.35-1.16) | 0.145 |
| >5.1 | 0.15 (0.07-0.33) | 0.000 | 0.62 (0.41-0.94) | 0.023 | 0.40 (0.20-0.78) | 0.008 |

\* Reference category

### Predictors of ADR

Univariate logistic regression analysis exploring predictors of acquired drug resistance is shown in Table F in Additional File 1. Being male, an age at diagnosis above 26 years old, being MSM (compared to being heterosexual), being born in Europe, being infected with subtype B and presenting logVL above 4.1 presented statistically significant results.

Multivariate analysis showed statistical significance for any ADR and NRTIs drug resistance for males, infected with subtype B and presenting viral loads above 4.1 (Table 4).

**Table 4.** Multiple regression analysis of factors associated with HIV acquired drug resistance

| Variable | Any ADR |  | NRTI ADR |  | NNRTI ADR |  | PI ADR |  |
| --- | --- | --- | --- | --- | --- | --- | --- | --- |
|  | OR (95%CI) | p | OR (95%CI) | p | OR (95%CI) | p | OR (95%CI) | p |
| **Gender** |  |  |  |  |  |  |  |  |
| Female * |  |  |  |  |  |  |  |  |
| Male | 1.44 (1.20-1.73) | 0.000 | 1.40 (1.18-1.67) | 0.000 | 1.15 (0.97-1.35) | 0.112 | 1.66 (1.35-2.05) | 0.000 |
| **Age at diagnosis** |  |  |  |  |  |  |  |  |
| 18-25* |  |  |  |  |  |  |  |  |
| 26-40 | 1.12 (0.73-1.73) | 0.602 | 1.13 (0.74-1.72) | 0.565 | 0.85 (0.57-1.27) | 0.420 | 1.71 (0.95-3.07) | 0.072 |
| 41-55 | 1.07 (0.69-1.67) | 0.764 | 1.11 (0.72-1.70) | 0.626 | 0.83 (0.55-1.25) | 0.375 | 1.76 (0.97-3.18) | 0.062 |
| >56 | 1.27 (0.76-2.12) | 0.367 | 1.37 (0.84-2.24) | 0.207 | 0.84 (0.53-1.35) | 0.481 | 2.65 (1.40-5.02) | 0.003 |
| **Subtypes** |  |  |  |  |  |  |  |  |
| B* |  |  |  |  |  |  |  |  |
| Non-B | 0.68 (0.57-0.80) | 0.000 | 0.61 (0.52-0.72) | 0.000 | 0.98 (0.84-1.14) | 0.789 | 0.64 (0.54-0.77) | 0.000 |
| **Viral Load (log<sub>10</sub> copies/ml)** |  |  |  |  |  |  |  |  |
| < 4.0* |  |  |  |  |  |  |  |  |
| 4.1 to 5.0 | 0.63 (0.52-0.76) | 0.000 | 0.60 (0.50-0.71) | 0.000 | 0.89 (0.75-1.05) | 0.165 | 0.85 (0.70-1.03) | 0.090 |
| >5.1 | 0.34 (0.27-0.42) | 0.000 | 0.29 (0.23-0.36) | 0.000 | 0.69 (0.56-0.85) | 0.001 | 0.50 (0.38-0.65) | 0.000 |

## Discussion

Our study showed that the estimated prevalence of TDR over time, among DN patients, increased between 2003 and 2017, and this increase was more pronounced between 2014 and 2017. While TDR to NRTIs and NNRTIs increased, TDR to PIs decreased me. We hypothesize that this increase could have different explanations. One hypothesis, that we have tested, is the implementation of the Treatment for All recommendations. The increasing trend for TDR is steeper since 2014, which could indicate a contribution of this public health measure to the increase of TDR. However, other factors could also be associated, namely the changing face of the epidemic, with an increasing proportion of MSMs among new diagnoses. This population is associated with faster onward transmission of HIV infection, which could potentiate transmission of TDR. Finally, the increasing mobility of populations and the presence in Portugal of a higher number of migrants from countries where TDR has been increasing in the last years could also explain for this trend [11]. While we would expect that this should be seen in the proportion of TDR in non-B subtypes and we don’t see it, we are exploring this in other independent analyses. Similarly to our study, a study of HIV drug resistance in a Canadian cohort showed an increase in TDR between 1996 and 2014, with a more prominent increase in the last years [12], which is consistent with our results. Some other studies, however, showed opposite results, with TDR prevalence declining or stabilizing over time [13–17]. A potential explanation for our discrepant results could be the origin of migrants living in Portugal, which come from Portuguese speaking Sub-Saharan African countries, where TDR has been increasing in the last years [18,19].

Strikingly, in the same time period (2001-2017) we found a significant declining trend in ADR, that span all drug classes. This trend has been consistently found in other studies. For example, in a multi-center European cohort (1997-2008) in Switzerland it was demonstrated that the majority of treated patients who initiated treatment in more recent years did not acquire drug resistance [20], as well as in another study in a large Canadian cohort (1996-2016) that showed that the prevalence of ADR has been declining for all drug categories [12]. Concordant results were also observed in an Italian cohort (2003-2012), in a German cohort (2001-2011), in Spain (1999-2005) and in Western Europe (1997-2008) [13,21–23]. Rocheleau et al (2017) have proposed that this declining trend of ADR is due to a combination of factors, that includes the increased efficacy of ARV regimens, readily accessible combination regimens and improved patient management [12].

The most frequently detected mutations in DN patients were K103NS conferring high-level resistance to NVP and EFV; and M41L, which reduces susceptibility to TDF and ABC when in combination with other NRTIs mutations. M184VI associated with high-level resistance to NRTIs also causes high-level reduced susceptibility to 3TC, FTC and ABC. L90M was the resistance mutation to PIs with highest prevalence. It causes reduced susceptibility to ATV and LPV when in combination with other PI-resistance mutations. While the transmission of K103NS is not that worrying given that NNRTIs are no longer recommended as first line treatment, M41L and M184IV have an impact to drugs recommended for first-line, so it is important to understand the impact of the transmission of these mutations and why its prevalence is increasing among DN patients (Figure 3). Furthermore, first line treatment recommendations are not used in many African countries, where regimens used for first line include drugs to which all these mutations cause resistance. It is also important to keep in mind that increasing mobility of populations can cause the spread and transmission of ARV drug resistance mutations in settings where other drugs are used.

For treated patients, the more prevalent mutations conferring resistance to NRTIs, were M184IV conferring high-level resistance to FTC and 3TC and also reduced susceptibility to ABC. K103NS was the most prevalent mutation conferring resistance to NNRTIs, specifically to NVP and EFV and L90M reducing susceptibility in combination with other PIs mutations, specifically to ATV and LPV. Importantly, overtime we observed an important reduction of specific PIs and NRTIs mutations, such as M46IL, L90M, M41L, L210W and M184IV. This could be associated with higher genetic barrier of boosted PIs and of dual NRTIs formulations (2014) [24].

According to the multivariate analysis performed in this study, the risk of TDR (and ADR) is significantly lower in patients infected with non-B subtypes compared to B subtype, as previously shown [25–28]. We hypothesize that this could be due to longer use of ARV therapy in developed countries, where this subtype is more prevalent, and to the circulation of this subtype in MSMs, where transmission chains are faster and therefore potentiate the transmission of TDR. However, there are features of the changing HIV-1 epidemic that could lead to an inversion of this pattern. The treatment for all, the increasing prevalence of non-B subtypes in developed countries and, particularly, the growing number of reports indicating transmission clusters of non-B subtypes in MSM could lead to an increase of TDR in non-B subtypes.

Our multivariate analysis also showed that patients with TDR and ADR tended to present lower levels of VL at the time of resistance testing, which is consistent with the findings of the Swiss cohort [29], that found the same results in a population of DN patients and patients non-respondent to first line regimens. It was also observed that lower VL of DN patients was associated with a higher level of patients presenting M184V and L90M mutation with statistical significance. Due to its high fitness cost [30], M184V quickly disappears after transmission in the absence of drug pressure [31]. These results are according with previous studies suggesting that higher viral load promote loss of mutations [29,30]. However, for K103NS no statistical association was found, which may be due to the differential fitness costs in relation to M184V, since K103NS are low-fitness-cost mutations that persist longer [32,33]. Other studies have shown that specific TDR mutations have an effect on the viral fitness that can have implications for its transmissibility [34,35].

We also found that the risk of ADR was significant higher in male patients, [13], in patients infected with subtype B and in patients with lower levels of VL at the time of resistance testing. ADR can be related to poor adherence, which has been shown to be a major determinant of virologic failure and emergence of drug resistant virus. Barriers to optimal adherence may originate from individual (biological, socio-cultural, behavioral), pharmacological, and societal factors; which make these findings more complex to interpret. Other studies also associated the development of ADR with levels of adherence in each era of therapy initiation [12,13].

### Conclusion

In summary, our study showed that while ADR is declining since 2001, TDR has been steadily increasing, reaching a worrying value of 13.1% in 2017. While declining ADR seems to be caused mainly by the increasing efficacy of ARV therapy, TDR seems to be mainly driven by determinants of the virus such as viral subtype and fitness of the virus in the presence of particular mutations. Our results highlight that It is urgent to develop public health programs to monitor levels and patterns of TDR in newly diagnosed patients.

## Data Availability

Data are available upon request

## Authors’ contributions

Conceptualization: MP and AA; Data curation: ID, SF, MP, AP and VP; Formal analysis: MP, AP, MM, PL, KT and AA; Funding acquisition: AA; Investigation: MP, VP, PG, AV, RC and PRSG; Methodology: MP, AP, VP and AA; Project administration: AA; Resources: PG, RC, AV and PRSG; Supervision: AA; Validation: MP and MM; Writing – original draft: MP and AA; Writing – review & editing: MP and AA.

### Acknowledgements

We would like to thank the patients and all the members of the Portuguese HIV-1 Resistance Study Group: Fátima Gonçalves^2^, Joaquim Cabanas^2^, Ana Patrícia Carvalho^2^, Inês Costa^2^, Kamal Mansinho^6^, Ana Cláudia Miranda^6^, Isabel Aldir^6^, Fernando Ventura^6^, Jaime Nina^6^, Fernando Borges^6^, Emília Valadas^7^, Manuela Doroana^7^, Francisco Antunes^7^, Nuno Marques^8^, Maria João Aleixo^8^, Maria João Águas^8^, Júlio Botas^8^, Teresa Branco^9^, Patrícia Pacheco^9^, Luís Duque^9^, José Vera^10^, Luís Tavares^10^, Inês Vaz Pinto^11^, José Poças^12^, Joana Sá^12^, António Diniz^13^, Margarida Serrado^13^, Ana Mineiro^13^, Flora Gomes^14^, Cristina Guerreiro^14^, Carlos Santos^15^, Domitília Faria^15^, Paula Fonseca^15^, Paula Proença^16^, Telo Faria^17^, Eugénio Teófilo^18^, Sofia Pinheiro^18^, Isabel Germano^19^, Umbelina Caixas^19^, Margarida Bentes Jesus^19^, Nancy Faria^20^, Ana Paula Reis^20^, Graça Amaro^21^, Fausto Roxo^21^, Ricardo Abreu^22^ and Isabel Neves^22^.

## Affiliations

6- Serviço de Doenças Infeciosas, Centro Hospitalar de Lisboa Ocidental, Hospital de Egas Moniz, Lisboa, Portugal

7- Serviço de Infeciologia, Hospital de Santa Maria, Centro Hospitalar de Lisboa Norte, Lisboa, Portugal

8- Serviço de Infeciologia, Hospital Garcia da Orta, Almada, Portugal

9- Serviço de Infeciologia, Hospital Dr. Fernando da Fonseca, Amadora, Portugal

10- Serviço de Medicina Interna, Centro Hospitalar do Barreiro-Montijo, Portugal

11- Unidade funcional VIH/SIDA, Hospital de Cascais, Cascais, Portugal

12- Serviço de Infeciologia, Centro Hospitalar de Setúbal, Setúbal, Portugal

13- Serviço de Pneumologia, Hospital Pulido Valente, Centro Hospitalar de Lisboa Norte, Lisboa, Portugal

14- Serviço de Obstetrícia - Maternidade Alfredo da Costa, Centro Hospitalar de Lisboa Central, Lisboa, Portugal

15- Serviço de Medicina Interna-Centro Hospitalar do Algarve, Hospital de Portimão, Portimão, Portugal

16- Serviço de Infeciologia-Centro Hospitalar do Algarve, Hospital de Faro, Faro, Portugal

17- Unidade Local de Saúde do Baixo Alentejo, Hospital José Joaquim Fernandes, Beja, Portugal

18- Serviço de Infeciologia, Hospital de Santo António dos Capuchos, Centro Hospitalar de Lisboa Central, Lisboa, Portugal

19- Serviço de Medicina 1.4, Centro Hospitalar de Lisboa Central, Hospital de São José, Lisboa, Portugal

20- Serviço de Doenças Infectocontagiosas, Hospital Central do Funchal, Madeira, Portugal 21- Serviço de Doenças Infeciosas, Hospital Distrital de Santarém, Santarém, Portugal

21- Serviço de Infeciologia, Unidade de Local de Saúde de Matosinhos, Hospital Pedro Hispano, Matosinhos, Portugal

## Supplementary files legend

**Supplementary File 1- Text and Tables**

This file contains an explained text with the methodology used to do the statistical analysis as well as the characterization of the Portuguese population included in this study.

**Table A-** Demographic and clinic patient characteristics, 2001-2017

**Table B**- Trends of transmitted drug resistance between 2006 and 2017, 2014 and 2017 and September 2014 and December 2017 and of acquired drug resistance between 2001 and 2017

**Table C**- Proportion of transmitted drug resistance and of acquired drug resistance for single, double and triple classes and for associations of drug classes between 2001 and 2017

**Table D-** Time trends of selected mutations with prevalence greater than 0.5% for drug-naïve patients and 5.0% for treated patients

**Table E**- Univariate analysis of factors associated with HIV transmitted drug resistance

**Table F**- Univariate analysis of factors associated with HIV acquired drug resistance

**Supplementary Figure 1:**
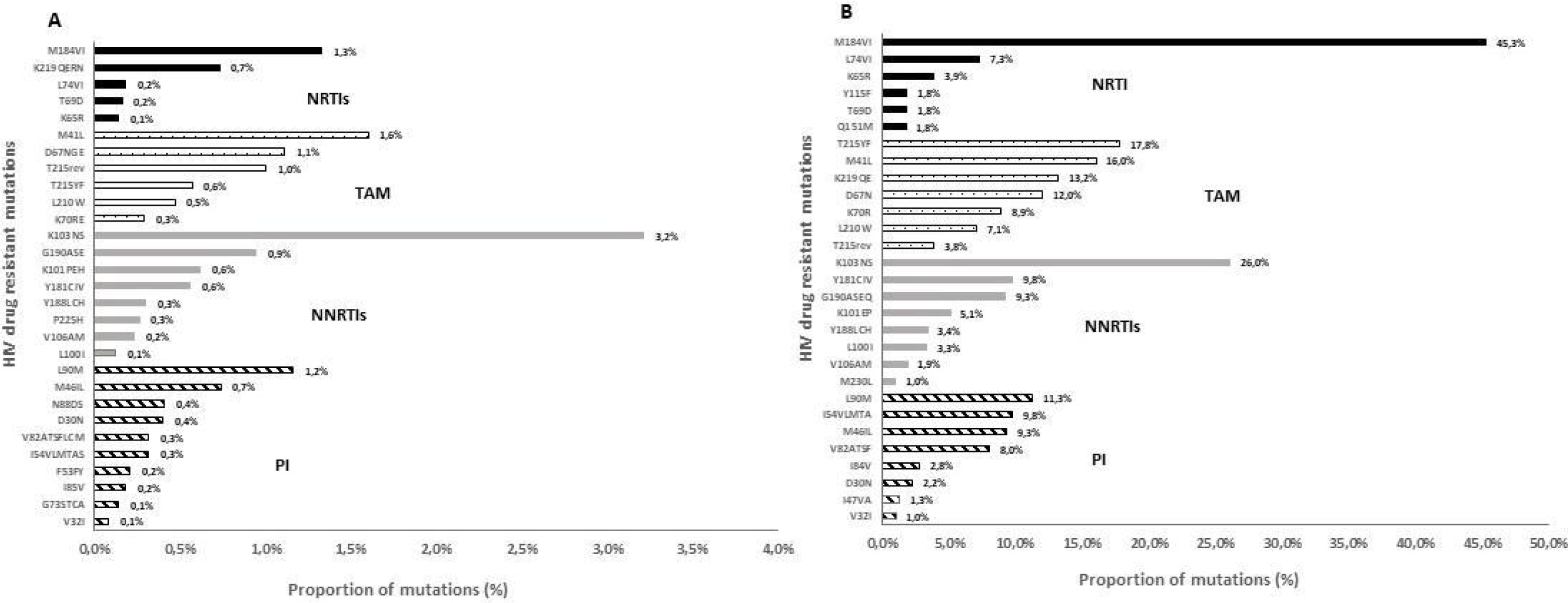
Proportion of resistance mutations in sequences (A) drug naïve patients and (B) treated patients between 2001 and 2017

